# Predicting the number of reported and unreported cases for the COVID-19 epidemics in China, South Korea, Italy, France, Germany and United Kingdom

**DOI:** 10.1101/2020.04.09.20058974

**Authors:** Z. Liu, P. Magal, G. Webb

## Abstract

We model the COVID-19 coronavirus epidemics in China, South Korea, Italy, France, Germany and United Kingdom. We use early reported case data to predict the cumulative number of reported cases to a final size in each country. The key features of our model are the timing of implementation of major public policies restricting social movement, the identification and isolation of unreported cases, and the impact of asymptomatic infectious cases.

## 1 Introduction

COVID-19 epidemics are currently a global crisis. Our goal in this paper is to develop a mathematical model to guide understanding of the dynamics of COVID-19 epidemics. In previous works [5, 6, 7], our team has developed differential equations models of COVID-19 epidemics. Our goal is to predict forward in time the future number of cases from early reported case data in regions throughout the world. Our models incorporate the following important elements of COVID-19 epidemics: (1) the number of asymptomatic infectious individuals (with no or very mild symptoms), (2) the number of symptomatic reported infectious individuals (with severe symptoms) and (3) the number of symptomatic unreported infectious individuals (with mild symptoms). With our models, we provide prediction of the final size of the asymptomatic infectious cases, the reported cases (with severe symptoms) and unreported cases (with mild symptoms). We will apply the model in this paper to the epidemics in China, South Korea, Italy, France, Germany, and the United Kingdom.

The significance of transmission by asymptomatic and mildly symptomatic individuals is of major importance. It is well established that newly infected patients, who are asymptomatic or have only mild symptoms, can transmit the virus [10, 11, 12, 15, 17, 19, 20]. In an early phase of the epidemic, the reported case data grows exponentially, which corresponds to a constant transmission rate. We assume that government measures and public awareness cause this early constant transmission rate to change to a time dependent exponentially decreasing rate. We identify this early constant transmission rate using a method developed in [5]. We then identify the time dependent exponentially decreasing transmission rate from reported case data, and project forward the time-line of the epidemic course. Our model is applicable to COVID-19 epidemics in any region with reported case data, which can be updated to higher accuracy with on-going day-by-day reported case data. We refer to [4] and [18] for related results.

The organisation of this paper is as follows. In Section 2 we present a general differential equations model for current COVID-19 epidemics, applicable to any region for which reported case data is available.

In Section 3 we provide a general method to parameterize and initialize the model. In Section 4 we apply the model to China, South Korea, Italy, France, Germany, and the United Kingdom. In Section 5 we discuss conclusions obtained from our analysis.

## 2 Model

The model consists of a mass action law epidemic model where the asymptomatic infectious and the unreported infectious can transmit the virus

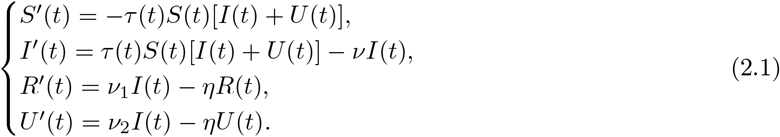

This system is supplemented by initial data

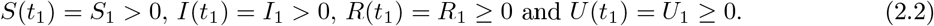

Here *t* ≥ *t*_1_ is time in days, *t*_1_ is the beginning date from which the number of reported symptomatic infectious individuals growths exponentially, *S*(*t*) is the number of individuals susceptible to infection at time *t, I*(*t*) is the number of asymptomatic infectious individuals at time *t, R*(*t*) is the number of reported symptomatic infectious individuals at time *t* (i.e. symptomatic infectious with sever symptoms), and *U* (*t*) is the number of unreported symptomatic infectious individuals at time *t* (i.e. symptomatic infectious with mild or no symptoms).

The transmission rate at time *t* is *τ* (*t*). Asymptomatic infectious individuals *I*(*t*) are infectious for an average period of 1*/ν* days. Reported symptomatic individuals *R*(*t*) are infectious for an average period of 1*/η* days, as are unreported symptomatic individuals *U* (*t*). We assume that reported symptomatic infectious individuals *R*(*t*) are reported and isolated immediately, and cause no further infections. The asymptomatic individuals *I*(*t*) can also be viewed as having a low-level symptomatic state. All infections are acquired from either *I*(*t*) or *U* (*t*) individuals. The fraction *f* of asymptomatic infectious become reported symptomatic infectious, and the fraction 1 − *f* become unreported symptomatic infectious. The rate asymptomatic infectious become reported symptomatic is *ν*_1_ = *f ν*, the rate asymptomatic infectious become unreported symptomatic is *ν*_2_ = (1 − *f*) *ν*, where *ν*_1_ + *ν*_2_ = *ν*. The cumulative number of reported cases at time *t* is obtained by using the following equation

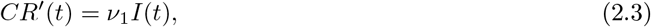

and the cumulative number of unreported at time *t* is given by the formula

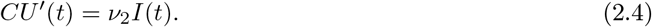

The parameters and initial conditions of the model are given in Table 1 and a flow diagram of the model is given in Figure 1.

**Table 1:**
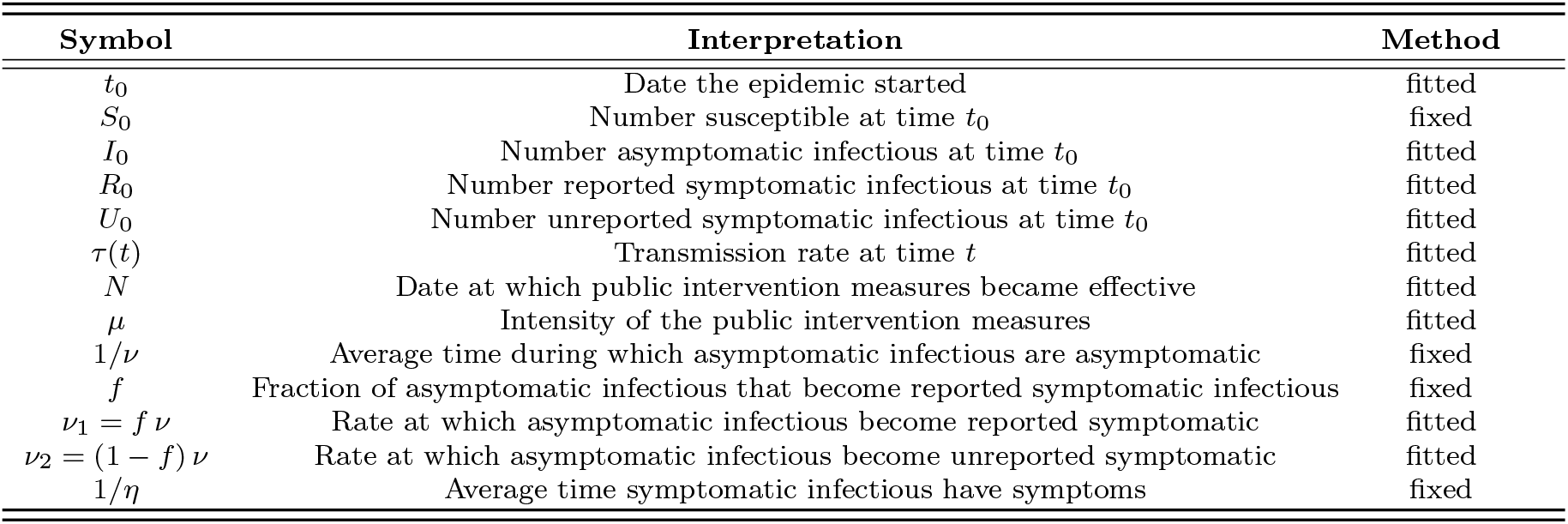
Parameters and initial conditions of the model.

| Symbol | Interpretation | Method |
| --- | --- | --- |
| $t_0$ | Date the epidemic started | fitted |
| $S_0$ | Number susceptible at time $t_0$ | fixed |
| $I_0$ | Number asymptomatic infectious at time $t_0$ | fitted |
| $R_0$ | Number reported symptomatic infectious at time $t_0$ | fitted |
| $U_0$ | Number unreported symptomatic infectious at time $t_0$ | fitted |
| $\tau(t)$ | Transmission rate at time $t$ | fitted |
| $N$ | Date at which public intervention measures became effective | fitted |
| $\mu$ | Intensity of the public intervention measures | fitted |
| $1/\nu$ | Average time during which asymptomatic infectious are asymptomatic | fixed |
| $f$ | Fraction of asymptomatic infectious that become reported symptomatic infectious | fixed |
| $\nu_1 = f\nu$ | Rate at which asymptomatic infectious become reported symptomatic | fitted |
| $\nu_2 = (1 - f)\nu$ | Rate at which asymptomatic infectious become unreported symptomatic | fitted |
| $1/\eta$ | Average time symptomatic infectious have symptoms | fixed |

**Figure 1:**
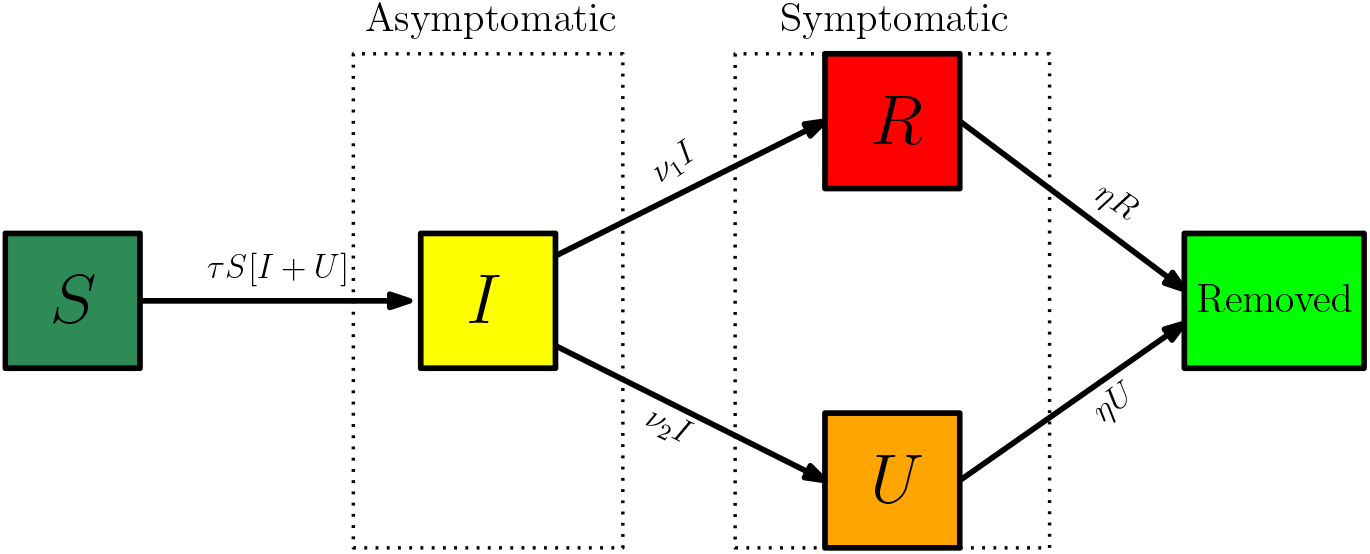
Compartments and flow chart of the model.

## 3 Method to estimate the parameters and initial conditions

We assume that 100 *× f* % of symptomatic infectious cases go unreported. The actual value of *f* is unknown and varies from country to country. We assume *η* = 1*/*7, which means that the average period of infectiousness of both unreported symptomatic infectious individuals and reported symptomatic infectious individuals is 7 days. We assume *ν* = 1*/*7, which means that the average period of infectiousness of asymptomatic infectious individuals is 7 days. These values can be modified as further epidemiological information becomes known.

At an early stage of the epidemic we assume that all the infected components of the system grow exponentially while the number of susceptible remains unchanged during a relatively short period of time *t* ∈ [*t*_1_, *t*_2_], which corresponds to a constant transmission rate *τ*_1_. Therefore, we will assume that

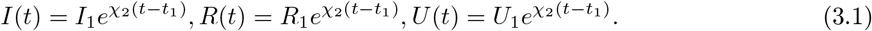

We deduce that the cumulative number of reported satisfies

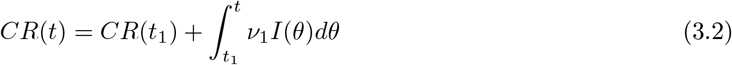

and by replacing *I*(*t*) by the exponential formula in (3.1), we have

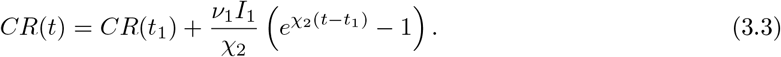

Hence it makes sense to assume that *CR*(*t*) − *CR*(*t*_1_) has the following form

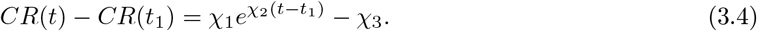

By identifying (3.3) and (3.4) we deduce that

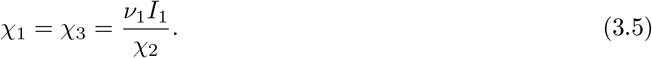

Moreover by assuming that the number of susceptible *S*(*t*) remains constant *S*_1_ on the time interval *t* ∈ [*t*_1_, *t*_2_], the *I*-equation, *R*-equation and *U* -equation of the model (2.1) become

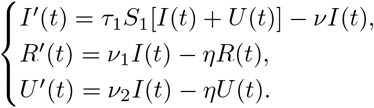

By using (3.1) we obtain

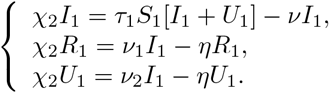

Computing further, we get

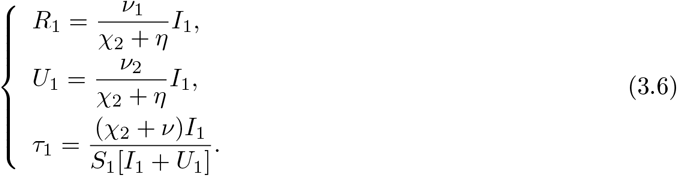

By (3.5) we deduce that

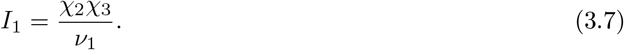

Finally we obtain

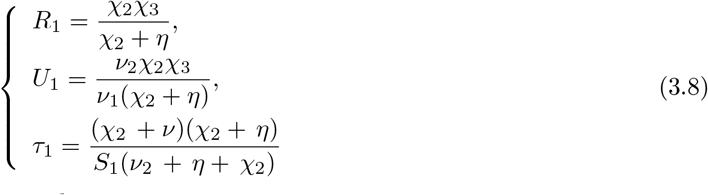

The value of the basic reproductive number is

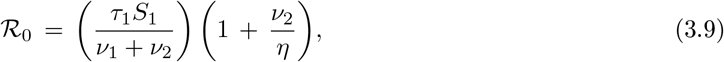

which was derived in [5].

During the exponential growth phase of the epidemic, *τ* (*t*) ≡ *τ*_1_ is constant. When strong government measures such as isolation, quarantine, and public closings are implemented, we use an exponential decrease for a time-dependent decreasing transmission rate *τ* (*t*) to incorporate these effects since the actual effects of these measures are complex. The formula for *τ* (*t*) during this phase is

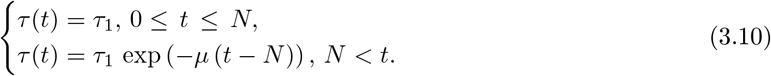

The date *N* and the value *µ* are chosen so that the cumulative reported cases in the numerical simulation of the epidemic aligns with the cumulative reported case data after day *N*, when the public measures take effect. In this way we are able to project forward the time-path of the epidemic after the government imposed public restrictions take effect.

The daily number of reported cases from the model can be obtained by computing the solution of the following equation:

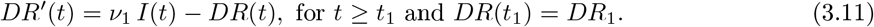

## 4 Predicting the cumulative number of cases

In this section we apply the method described in Section 3 to the data of confirmed cases in China, South Korea, Italy, France, Germany, and the United Kingdom. We predict the time evolution of a COVID-19 epidemic through its phases, we identify epidemic turning points by using the data for the cumulative number of reported cases and the daily number of reported cases, and we project the epidemic final sizes.

As we can see in Figure 2, a major difficulty for the predictions from the reported case data, is to determine the date intervals for exponential growth phase of the epidemic. For South Korea, the date intervals for exponential growth phase of the epidemic is relatively clear from February 25 to March 1. This is because the measures imposed for contact tracing and social distancing were very strong. For Italy, the interval is more difficult to ascertain. We choose March 1 to March 10, but these values may change as more data of reported cases becomes available. For France, Germany, and the United Kingdom, the interval is very difficult to know with the present data. Again, future reported case data, will clarify these intervals. In general, the dates for the exponential growth interval and the date *N*, are key elements of our model, and they depend strongly on the implementation of social distancing measures. If these measures are implemented gradually, then the difficulty is increased.

**Figure 2:**
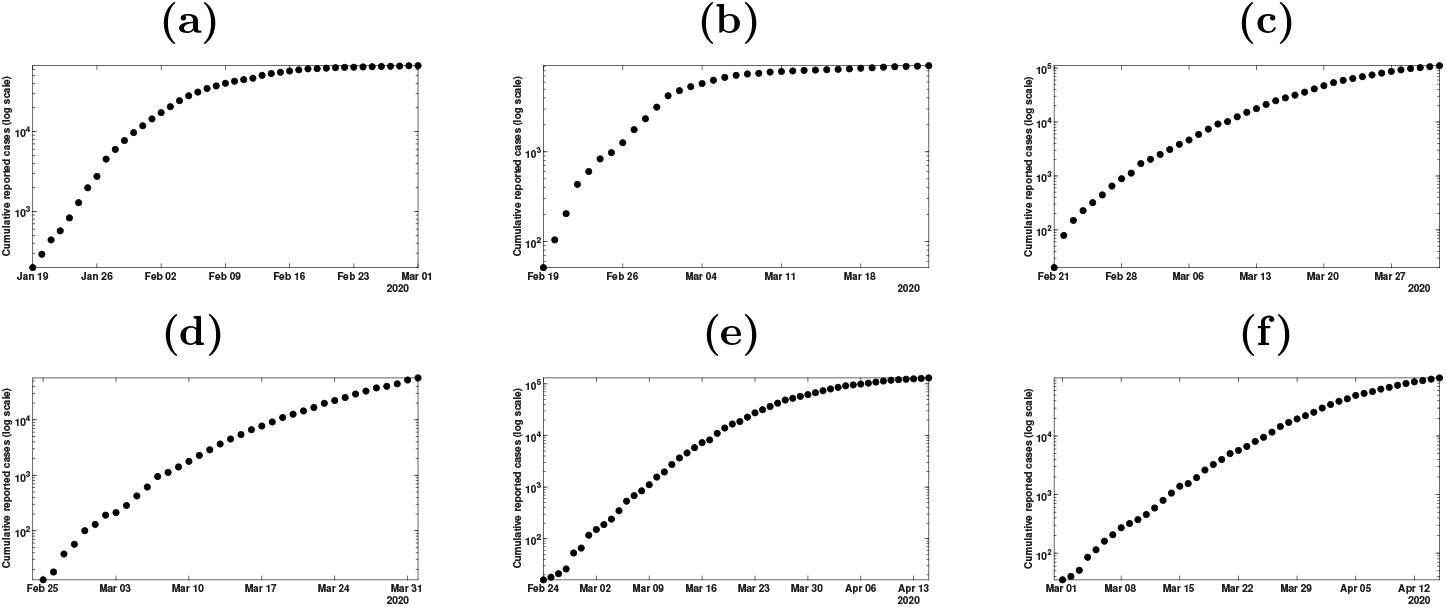
In this figure we plot the cumulative number cases (with log scale) at the early beginning of the epidemic for (a) mainland China; (b) South Korea; (c) Italy; (d) France; (e) Germany; (f) UK.

Another difficulty in applying our model is how to fix the values of the parameters *ν, f, η, µ, N*. We take *η* = 1*/*7, which means that the average period of infectiousness of both unreported symptomatic infectious individuals and reported symptomatic infectious individuals is 7 days, *ν* = 1*/*7, which means that the average period of infectiousness of asymptomatic infectious individuals is 7 days. These values follow from the medical and epidemiological information and can be modified as further epidemiological information becomes known. *N* is around the time point of the implementation of the national prevention and control measures. The values of *f* is unknown, but information about the level of testing relates to the value of *f*. A decreased value of *f* corresponds to a greater final size of the epidemic. The increased testing can increase the value of *f*. Mortality can also be used as a reference to estimate the value of *f*. High mortality indicates high unreported ratio. In fact, from the values of *f, N* and *µ*, we can also obtain some information of the actual effects of these measures of testing, quarantining and isolation implemented by the governments in these countries.

The principle of our method is the following. By using an exponential best fit method we obtain a best fit of (3.4) to the data over a time [*t*_1_, *t*_2_] and we derive the parameters *χ*_1_ and *χ*_2_. The values of *I*_1_ *U*_1_, *R*_1_ and *τ*_1_ are obtained by using (3.7)-(3.8). Next we fix *N* (first day of public intervention) to some value and we obtain *µ* by trying to get the best fit to the data.

In the method the uncertainty in our prediction is due to the fact that several sets of parameters (*t*_1_, *t*_2_, *N, f*) may give a good fit to the data. As a consequence, at the early stage of the epidemics (in particular before the turning point) the outcome of our method can be very different from one set of parameters to another. We try to solve this uncertainty problem by using several choices of the period to fit an exponential growth of the data to determine *χ*_1_ and *χ*_2_ and several choices for the first day of intervention *N*. So in this section, we vary the time interval [*t*_1_, *t*_2_], during which we use the data to obtain *χ*_1_ and *χ*_2_ by using an exponential fit. In the simulations below, the first day *t*_1_ and the last day *t*_2_ vary such that

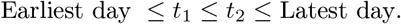

We also vary the first day of public intervention

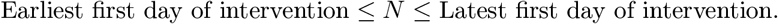

We vary *f* between 0.1 to 0.9. For each (*t*_1_, *t*_2_, *ν, f, η, µ, N*) we evaluate *µ* to obtain the best fit of the model to the data. We use the mean absolute deviation as the distance to data to evaluate the best fit to the data. We obtain a large number of best fit depending on (*t*_1_, *t*_2_, *ν, f, η, µ, N*) and we plot smallest mean absolute deviation MAD_min_. Then we plot all the best fit graphs with mean absolute deviation between MAD_min_ and MAD_min_ + 40.

## 5 Numerical simulations

The numerical simulations are presented in the chronological order of appearance for six countries (China, South Korea, Italy, France, Germany and United Kingdom).

### 5.1 Predicting the number of cases for mainland China

Before February 11, the data was based on confirmed testing. From February 11 to February 15, the data included cases that were not tested for the virus, but were clinically diagnosed. There were 17,409 such cases from February 11 to February 15 and there is a gap in the reported case data on February 11. The data from February 11 to February 15 specified both types of reported cases (i.e. the case tested for virus and the case clinically diagnosed). From February 16, the data did not separate the two types of reporting, but reported the sum of both types. We subtracted 17,409 cases (i.e. the new clinically diagnosed patient on Feb. 11) from the cumulative reported cases after February 15 to obtain the cumulative reported cases based only on confirmed testing after February 15.

In Figure 3, the predictions are improving with time. We observe that if we apply our method too early as in Figure 3 (a) and (b) the predictions vary a lot. But once we pass the turning point as in Figure 3 (c) and (d) the predictions are becoming pretty good and do not vary too much until the end of the epidemic.

**Figure 3:**
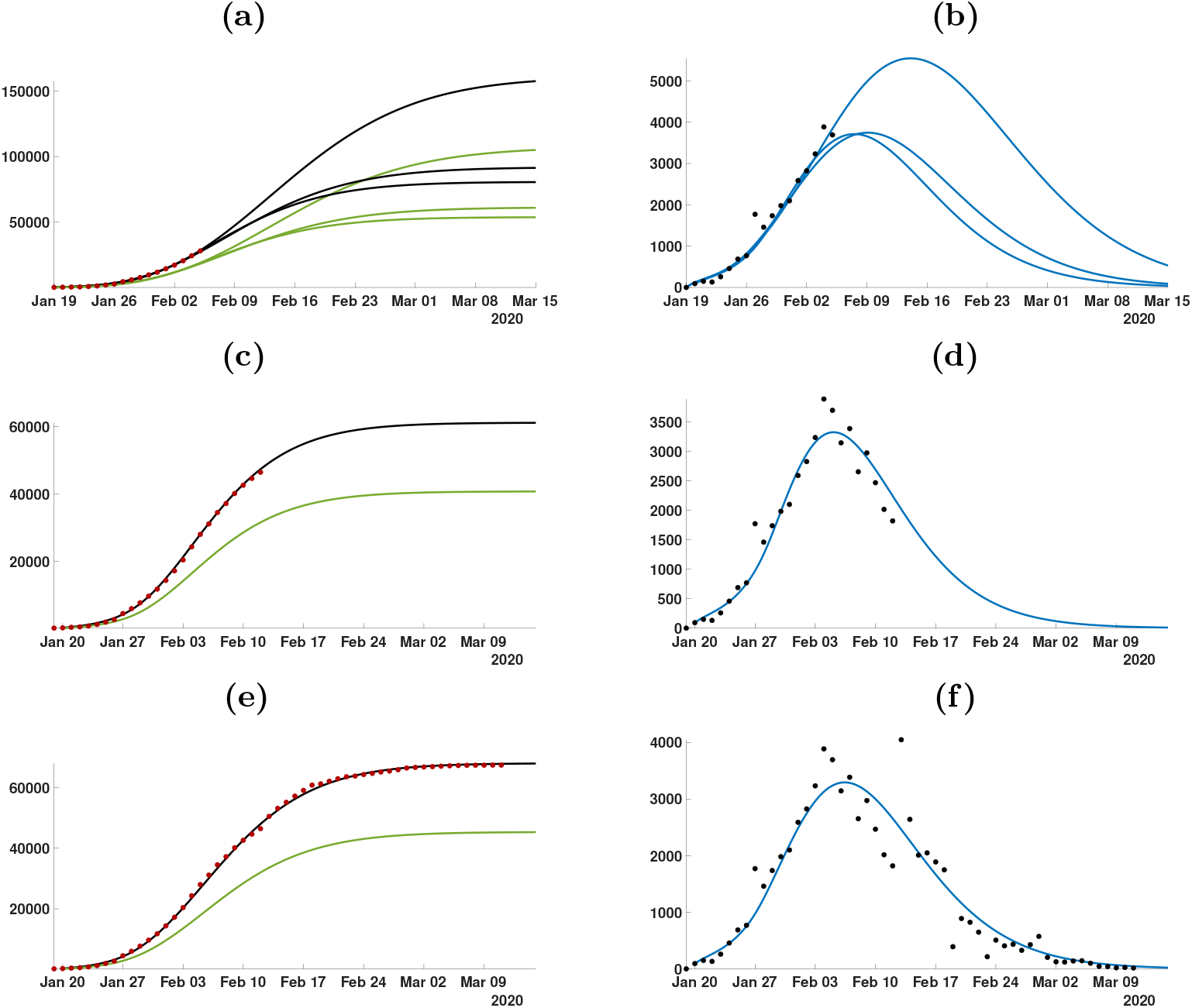
In this figure we plot the cumulative number of cases of the left hand side and the daily number of cases on the right hand side. In (a) and (b) we use the data until February 5. In (c) and (d) we use the data until February 12. In (e) and (f) we use the data until March 11. The best is obtained for t_1_ = January 20, t_2_ = January 30, 1/ν = 5 days, 1/η = 6 days, f = 0.6, µ = 0.1 and N = January 27. The smallest mean absolute deviation MAD_min_ is 475.

### 5.2 Predicting the number of cases for South Korea

The first confirmed case of the pandemic of COVID-19 in South Korea was announced on January 20, 2020. During the following four weeks (January 20 - February 17), South Korea monitored the potential spread of COVID-19 from existing confirmed patients, by using technological resources, such as tracking credit card use and checking CCTV footage of confirmed patients. On February 18, South Korea confirmed its 31^*st*^ case in Daegu. This 31^*st*^ patient had continued to go to gatherings of Shincheonji for days after showing symptoms. On February 19, the number of confirmed cases increased by 20 and on February 20 by 70, with a total of 346 confirmed cases by February 21, 2020, according to the Korean Centers for Disease Control (KCDC).

On February 20, the streets of Daegu were empty in reaction to the Shincheonji outbreak. On February 23, the government raised the coronavirus alert to ‘highest level’, and increased testing and contact tracing. Extensive measures were taken to screen the population for the virus, to isolate any infected people, and to quarantine those who were in contact with them. These rapid and extensive measures taken by the South Korean government have been judged successful in limiting the spread of the outbreak, without using the extreme measure of quarantining entire cities.

In Figure 4, the predictions are improving with time. As in Figure 3, when we apply our method too early as in Figure 4 (a) and (b) the result is very uncertain.

**Figure 4:**
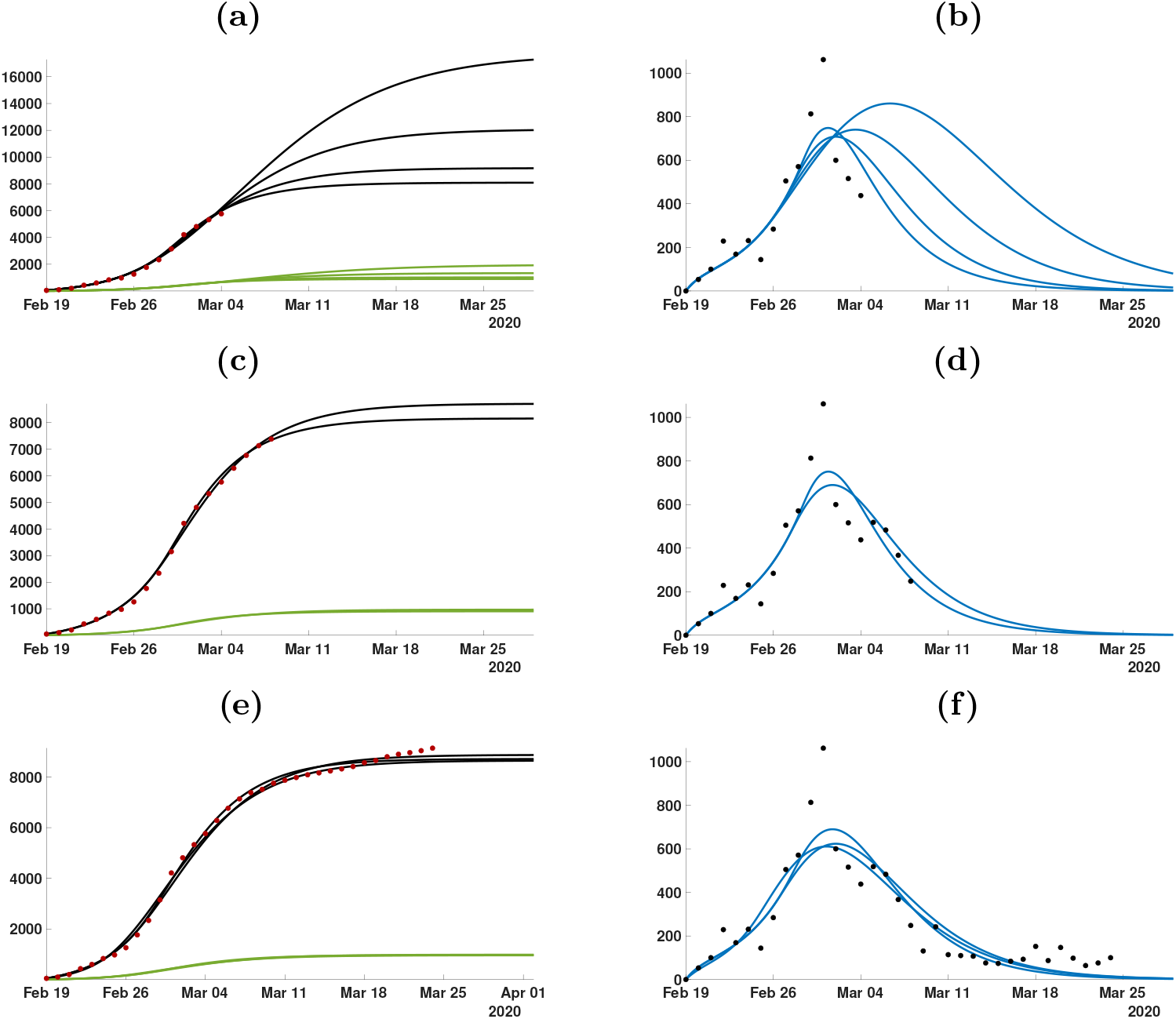
In this figure we plot the cumulative number of cases of the left hand side and the daily number of cases on the right hand side. In (a) and (b) we use the data until March 4. In (c) and (d) we use the data until March 8. (e) and (f) we use the data until March 24. The best is obtained for t_1_ = February 19, t_2_ = February 24, 1/ν = 4 days, 1/η = 16 days, f = 0.9, µ = 0.26 and N = February 27. The smallest mean absolute deviation MAD_min_ is 144.

**Figure 5:**
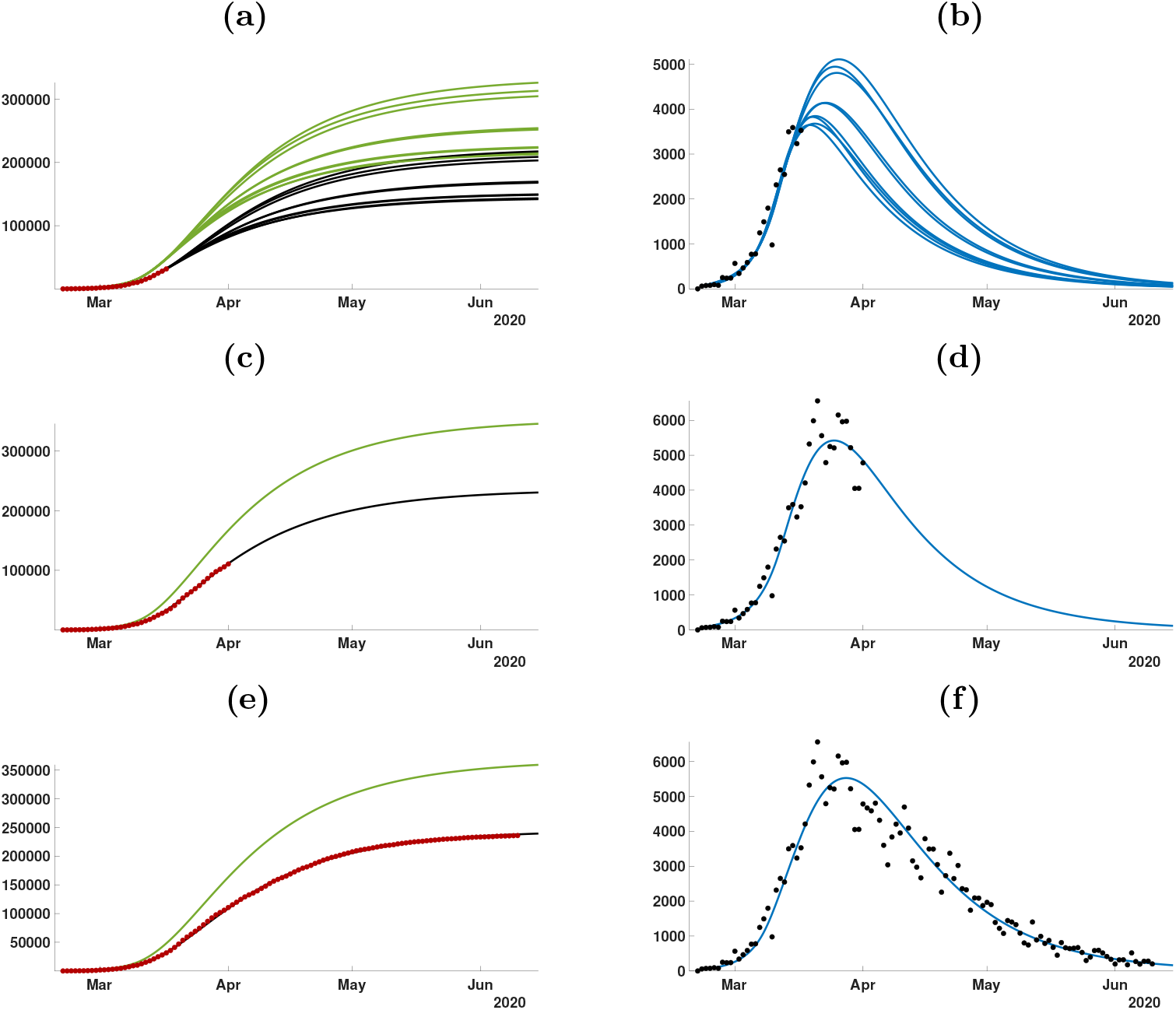
In this figure we plot the cumulative number of cases of the left hand side and the daily number of cases on the right hand side. In (a) and (b) we use the data until March 3. In (c) and (d) we use the data until March 10. (e) and (f) we use the data until March 24. The best is obtained for t_1_ = February 23, t_2_ = March 10, 1/ν = 23 days, 1/η = 14 days, f = 0.4, µ = 0.11 and N = March 10. The smallest mean absolute deviation MAD_min_ is 1129.

### 5.3 Predicting the number of cases for Italy

On January 31, the first two cases of COVID-19 were confirmed in Rome. On February 22, the government announced a series of isolation measures, such as a new decree imposing the quarantine of more than 50,000 people from 11 different municipalities in Northern Italy. On March 1, nationwide measures began to be implemented. On March 8, Prime Minister Giuseppe Conte extended the quarantine lockdown to cover all the region of Lombardy and 14 other northern provinces. On March 10, Prime Minister Conte increased the quarantine lockdown to cover all of Italy, including travel restrictions, a ban on public gatherings, and a shut down of all commercial and retail businesses except those providing essential services, such as grocery shops and pharmacies. On March 20, the Italian Ministry of Health ordered tighter regulations on free movement. The epidemic in Italy can be divided into the exponential growth phase before February 22. It is reasonable to take *N* between February 22 and March 10, because the government gradually strengthened control measures to the whole country during the period February 22 to March 10.

### 5.4 Predicting the number of cases for France

On January 24, the first COVID-19 case was confirmed in Bordeaux, France. The epidemic in France can be divided into the exponential growth phase before March 14. It is reasonable to take *N* between March 14 and March 17. The reasons are as follows: (1) On March 14, many cultural institutions in the Paris region announced their closure, such as the Louvre, the Eiffel Tower, and institutions in other provinces such as the Chateau de Montsoreau Museum of Contemporary Art; (2) French President Emanuel Macron delivered a speech on March 16, announcing that France entered a “state of war”, and closure measures would be imposed throughout all of France. On March 17, the “ban on foot” was officially launched.

### 5.5 Predicting the number of cases for Germany

A COVID-19 case was confirmed to have been transmitted to Germany on January 27, 2020. The epidemic in Germany can be divided into the exponential growth phase before March 13. It is reasonable to take *N* between March 13 and March 15, for the following reasons: (1) On March 13, 14 of the 16 German federal states closed their schools and nurseries; (2) On March 14, several federal states widened their measures to limit public activities. For example, Berlin, Schleswig-Holstein, and Saarland closed bars and other leisure venues. Cologne forbade all public events in the city center.

### 5.6 Predicting the number of cases for United Kingdom

The United Kingdom was slow to implement public distancing measures. On 19 March, the government introduced the Coronavirus Act 2020, which gave discretionary emergency powers in restricting social care facilities, schools, police, the Border Force, local councils, and other public functions. The act went into effect on 25 March 2020. Closures to pubs, restaurants and indoor sports and leisure facilities were imposed by the Health Protection Regulations on March 21.

## 6 Conclusions

We have applied a method developed in [5] and [6] to predict the evolution of a COVID-19 epidemic based on reported case data in that region. Our method uses early data, when the epidemic is in its exponential growth phase, corresponding to a constant transmission rate. In [5] we demonstrated a method to identify this constant transmission rate. When public measures are begun in order to ameliorate the epidemic, a new phase begins, which we model with a time-dependent exponentially decreasing transmission rate in [6]. In [6] we applied this method to mainland China, and demonstrated the ability of our model to predict the forward time-line of the epidemic. In Figure 4 in [6], we showed how the prediction unfolded week by week, with increasing agreement with reported case data, in mainland China.

**Figure 6:**
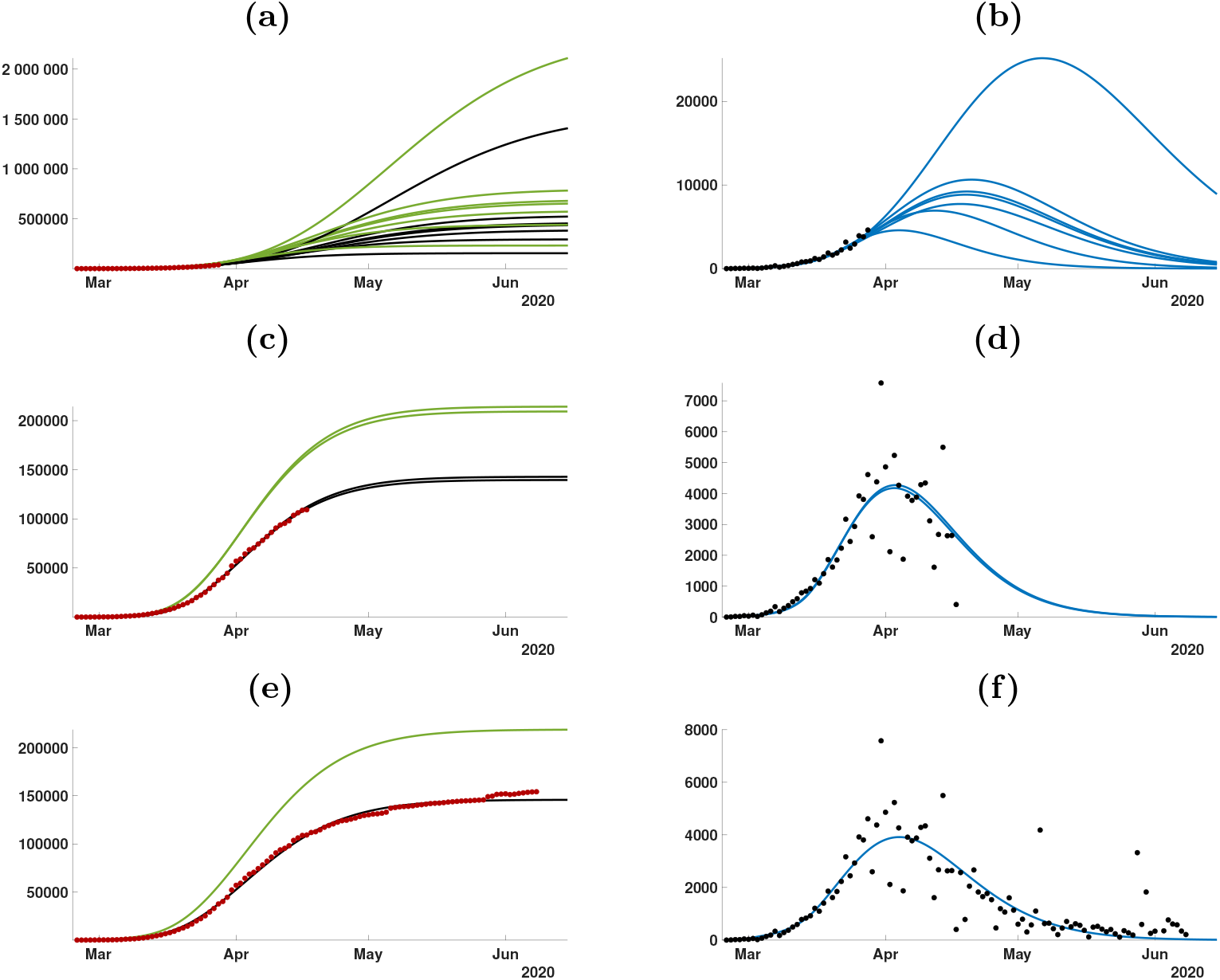
In this figure we plot the cumulative number of cases of the left hand side and the daily number of cases on the right hand side. In (a) and (b) we use the data until March 17. In (c) and (d) we use the data until April 1. (e) and (f) we use the data until June 10. The best is obtained for t_1_ = March 1, t_2_ = March 20, 1/ν = 6 days, 1/η = 15 days, f = 0.4, µ = 0.05 and N = March 15. The smallest mean absolute deviation MAD_min_ is 2347.

**Figure 7:**
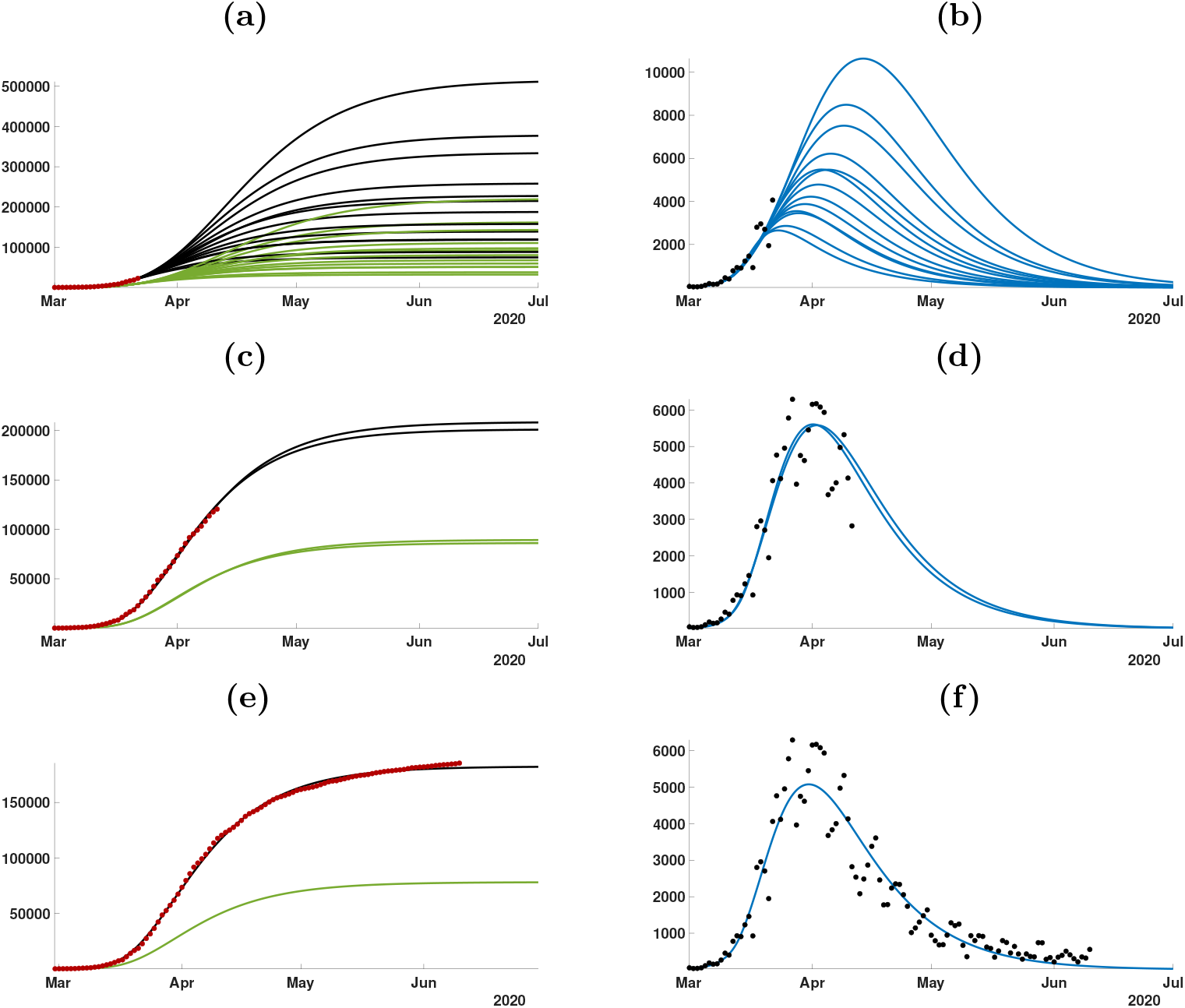
In this figure we plot the cumulative number of cases of the left hand side and the daily number of cases on the right hand side. In (a) and (b) we use the data until March 22. In (c) and (d) we use the data until April 11. (e) and (f) we use the data until June 10. The best is obtained for t_1_ = February 24, t_2_ = March 11, 1/ν = 15 days, 1/η = 14 days, f = 0.7, µ = 0.1 and N = March 13. The smallest mean absolute deviation MAD_min_ is 1368.

**Figure 8:**
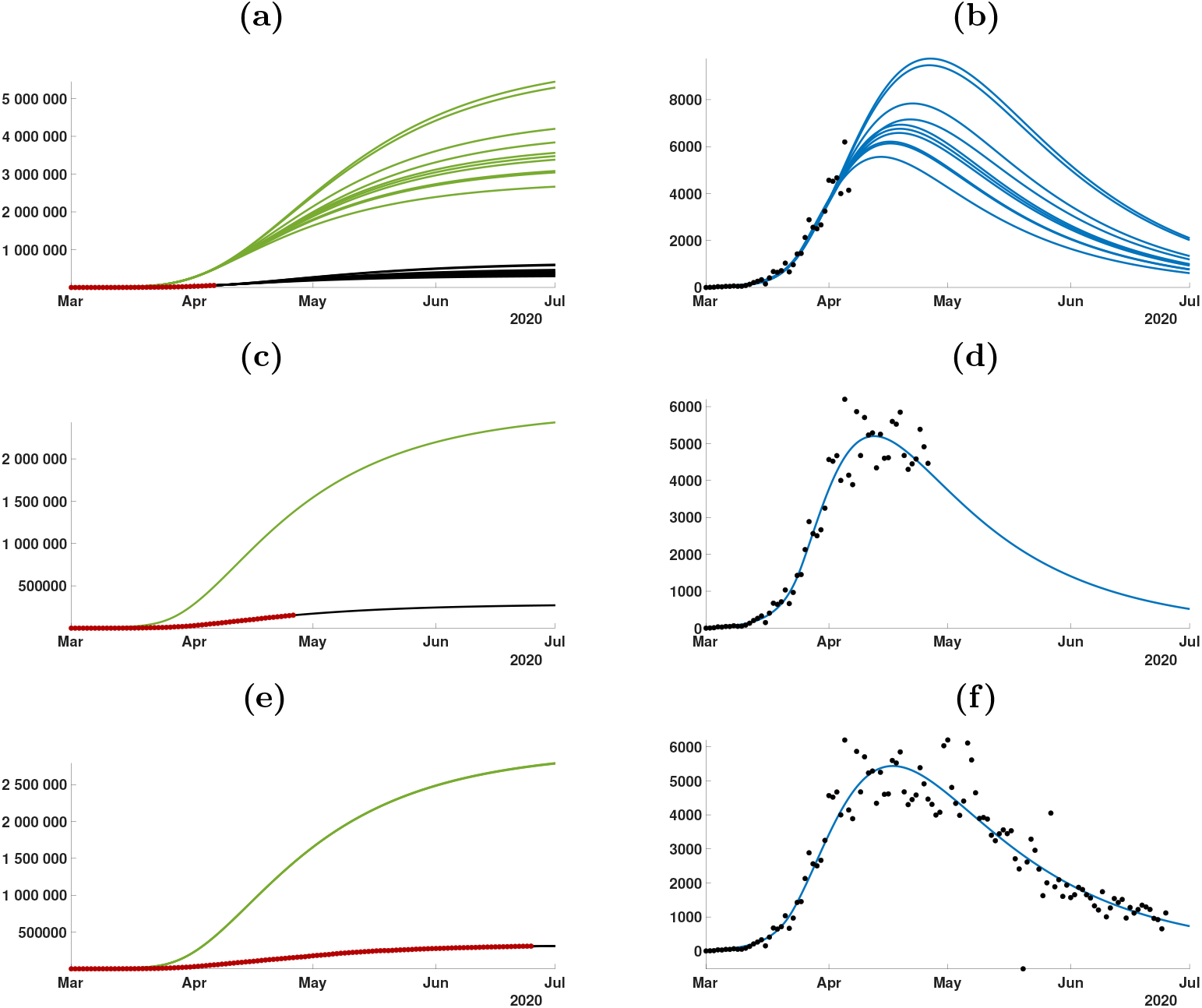
In this figure we plot the cumulative number of cases of the left hand side and the daily number of cases on the right hand side. In (a) and (b) we use the data until April 6. In (c) and (d) we use the data until April 26. (e) and (f) we use the data until June 25. The best is obtained for t_1_ = March 4, t_2_ = March 26, 1/ν = 30 days, 1/η = 31 days, f = 0.1, µ = 0.08 and N = March 22. The smallest mean absolute deviation MAD_min_ is 1248.

In Table 2 we summarized the parameters giving the best fit to the data.

**Table 2:** The parameters χ_1_, χ_2_, χ_3_ are estimated by using the data to fit χ_1_ exp(χ_2_ t) to the cumulative reported cases data CR(t) + χ_3_ (χ_3_ is chosen to minimize the error of the fit). For each country the best fit of the data over the all period is obtained by choosing: (1) t_1_ = January 27 and t_2_ = January 30 for mainland China; (2) t_1_ = February 19 and t_2_ = February 24 for South Korea; (3) t_1_ = February 23 and t_2_ = March 10 for Italy; (4) t_1_ = March 1 and t_2_ = March 20 for France; (5) t_1_ = February 24 and t_2_ = March 11 for Germany; (6) t_1_ = March 4 and t_2_ = March 26 for United Kingdom. The parameters ν = 1/7 and η = 1/7. The values of I_1_, R_1_, U_1_, τ_1_ are obtained by using (3.7), (3.8), and (3.4).

| Country | $\chi_1$ | $\chi_2$ | $\chi_3$ | $t_1$ | $\mu$ | $N$ | $S_1$ | $\tau_1$ |
| --- | --- | --- | --- | --- | --- | --- | --- | --- |
| China | 5.07 | 0.25 | 800.47 | Jan. 20 | 0.1 | Jan. 27 | $1.40005 \times 10^9$ | $2.73 \times 10^{-10}$ |
| South Korea | 3.529 | 0.24 | 319.67 | Feb. 19 | 0.26 | Feb. 27 | $51.47 \times 10^6$ | $8.82 \times 10^{-9}$ |
| Italy | 9.52 | 0.18 | 583.47 | Feb. 23 | 0.11 | Mar. 10 | $60.48 \times 10^6$ | $3.36 \times 10^{-9}$ |
| France | 4.86 | 0.16 | 717.7 | March 1 | 0.05 | Mar. 15 | $66.99 \times 10^6$ | $3.41 \times 10^{-9}$ |
| Germany | 0.0389 | 0.27 | 26.07 | Feb. 24 | 0.1 | Mar. 13 | $82.79 \times 10^6$ | $4.76 \times 10^{-9}$ |
| United Kingdom | 121.83 | 0.17 | 287.31 | Mar. 4 | 0.08 | Mar. 22 | $66.44 \times 10^6$ | $2.75 \times 10^{-9}$ |

In this work we use data for the cumulative number of reported cases and the daily number of reported cases for China, South Korea, Italy, France, Germany, and the United Kingdom. With this data, we project the future number of cases, both reported and unreported, in each country. For each country we observe that before the turning point the prediction is very uncertain and several of parameter values of *f ν η* may fit very well the data before the turning point. However, if we use enough reported case data, such as data up to the turning point, the prediction is pretty good and certain until the end of the epidemic.

A major difficulty for the predictions from the reported cases data, is to determine the date intervals of exponential growth phase. From the Figure 2 (a) this period is clear for mainland China (Jan 19-27). But it is much more difficult to decide for South-Korea (see Figure 2 (a)). By applying our method to the data for South Korea, we find the interval going from February 19 to February 24. This is because the measures imposed for contact tracing and social distancing were very strong. For Italy, the interval is difficult to ascertain (see Figure 2 (c)). We choose February 23 to March 10, but these values may change as more data of reported cases becomes available. For France, Germany, and the United Kingdom, the interval are also difficult to know with present data. Again, future reported case data, will clarify these intervals.

In the case of South Korea, the peak of the epidemic occurred approximately on February 29. In Figure 4, we see that our model agrees very well the data for South Korea. Accordingly to our model, the daily number of cases reaches a maximum of approximately 700 cases near the turning point February 29. Compared to South Korea, the public interventions in Italy, France, and Germany were relatively late. The peak of the epidemic occurs in Italy around March 14, and the peak of the maximum daily number of cases in our simulation is approximately 6 000, which agrees well with the daily reported cases data for Italy. For France, Germany, and the United Kingdom, the number of daily reported cases may still be rising. Our simulations captures these increasing values for exponential growth phase, but the advance to the next phase for both countries requires advanced data.

In general, the dates for the exponential growth interval, and the date *N* for the start of the new phase after the exponential growth interval, are key elements of our model, and they depend strongly on the implementation of social distancing measures. If these measures are implemented gradually, then the difficulty is increased. In future work we will allow for a series of *N* and *µ* values to account for a more complex implementation of measures that use a time-dependent transmission function *τ* (*t*) depending on these values.

Another difficulty in applying our model is the assumption of the fraction *f* of total cases reported. The value of *f* is unknown, but we vary *f* between 0.1 to 0.9. In future work, we will allow a time-dependent value for *f*, because in many applications the fraction of reported cases changes over the course of the epidemic. Our model incorporates social distancing measures through the time dependent transmission rate *τ* (*t*). It is evident that these measures should start as early as possible, and should be as strong as possible. The consequences of late public interventions may have severe consequences for the epidemic outcome. In future work we will apply our methods to other countries and regions within countries in an effort to better understand the evolution of COVID-19 epidemics.

In Table 3 we summarized the parameters giving the best fit to the data.

**Table 3:** In this table, we summarize the values for 1/ν (mean duration in days of the asymptomatic phase before symptoms onset), 1/η (mean duration in days of the symptomatic phase), and f (the fraction of reported) obtained for the best fits to the cumulative reported data.

| Country | $1/\nu$ | $1/\eta$ | $f$ |
| --- | --- | --- | --- |
| China | 5 days | 6 days | 0.6 |
| South Korea | 4 days | 16 days | 0.9 |
| Italy | 23 days | 14 days | 0.4 |
| France | 6 days | 15 days | 0.4 |
| Germany | 15 days | 14 days | 0.7 |
| United Kingdom | 30 days | 31 days | 0.1 |

The values of 1*/ν* (mean duration in days of the asymptomatic phase before symptoms onset) and 1*/η* (mean duration in days of the symptomatic phase) are very uncertain. Reported values have a wide range of variability. In an early report, a value of the asymptomatic phase was given as 5 days in [2]. WHO reported that the asymptomatic period is on average 5 − 6 days, but can be up to 14 days [23]. In recent reports, patients were reported asymptomatic 4 -17 days after admission to hospitals [8], and 9.5 days [3]. The Centers for Disease Control reported an average value of 6 days for asymptomatic transmission [1]. The period of median viral shredding was reported as 19 days pre-symptomatic patients and 14 days for symptomatic patients [8], and 22.6 days in pre-symptomatic patients and 25.2 days in symptomatic patients [13], although viral shedding does not necessarily correlate to infectiousness.

In this work many questions need future considerations. First the data are questionable and may induce a bias in the estimation of parameters. Probably the most significant effect on the data is induced by the daily fluctuations of the number of tests. Such a large fluctuation has been observed in China and other countries when the medical doctors were trying to adapt the number of tests to the number of symptomatic patients. Such a phenomenon may induce a large variation in the number of reported cases. In order to improve our understanding of the real situation from the cumulative reported cases, we can improve the model in several ways. For example we can introduce age groups (see [14, 9]) or we may examine big cities and their interconnections (see [21]). The method used to fit the data in the present paper has been extended and used successfully for age groups in Japan [9]. Such a method is general and may probably be applied to other situations (patch models etc …). In future work, we can also incorporate the number of tests (see [16]) which may significantly improve our understanding of COVID-19 epidemics.

## Data Availability

Data are available from WHO or Wikipedia

